# Cuticular Drusen Associated Photoreceptor and RPE Optical Property Perturbation Revealed by Adaptive Optics Scanning Laser Ophthalmoscopy

**DOI:** 10.64898/2026.01.15.26343733

**Authors:** Xiaolin Wang, Sujin Hoshi, Shin Kadomoto, Ruixue Liu, Michael Ip, David Sarraf, Srinivas R Sadda, Yuhua Zhang

## Abstract

**Purpose:** To characterize microscopic alteration of photoreceptors and RPE surrounding cuticular drusen in age-related macular degeneration (AMD) using multimodal imaging, including high resolution adaptive optics scanning laser ophthalmoscopy (AOSLO).

**Methods:** Eyes with early to intermediate AMD and predominantly cuticular drusen underwent color fundus photography, infrared reflectance, fundus autofluorescence, optical coherence tomography (OCT), and AOSLO. Cuticular drusen were identified using multimodal imaging and classified into three OCT-defined phenotypes. Cone photoreceptor reflectivity was assessed on AOSLO. A subset of eyes underwent longitudinal AOSLO and OCT imaging.

**Result:** Nineteen eyes from 12 subjects aged 70.3 ± 5.8 years were studied. Six eyes had longitudinal follow-up imaging. A total of 3177 cuticular drusen were evaluated and classified into 3 types based on cross sectional OCT imaging. AOSLO revealed corresponding phenotype-dependent cone reflectivity alterations associated with the 3 types of cuticular drusen. Type 1: Maintained cone reflectivity overlying the drusen on a hyporeflective background. Type 2: Cone reflectivity loss overlying the cuticular drusen. Type 3: Cones are predominantly not visible over the cuticular drusen. Lesion diameters were 52.62 ± 9.38 µm (Type 1), 71.88 ± 12.39 µm (Type 2), and 124.72 ± 20.94 µm (Type 3). All lesions were accompanied by hypertransmission in the choroid on OCT. Longitudinal imaging showed that localized outer retinal reflectivity reduction on AOSLO preceded the detection of new cuticular drusen on OCT.

**Conclusions:** Cellular-resolution multimodal imaging demonstrates progressive, phenotype-specific disruption of the photoreceptor–RPE complex associated with cuticular drusen in AMD. Early AOSLO-detected reflectivity changes preceding OCT-visible lesions highlight the sensitivity of adaptive optics imaging for identifying early outer retinal alterations and for advancing understanding of the biogenesis of cuticular drusen.

## Introduction

Age-related macular degeneration (AMD) is a multifactorial disease of the photoreceptor support system, including the RPE and inner choroidal vasculature, ultimately leading to photoreceptor dysfunction and vision loss.^1^ Although the initiating mechanisms underlying this outer retina–RPE–choroidal degeneration remain incompletely understood, one of the earliest detectable abnormalities is the accumulation of pathologic extracellular deposits, collectively termed drusen, between the basal lamina of the RPE and the inner collagenous layer of Bruch’s membrane (BrM).^2–6^ Cuticular drusen constitute a distinct subtype distinguished by characteristic morphology and clinical associations.^7–20^

Traditionally, cuticular drusen were characterized by small, round, yellow nodules on color fundus photography (CFP) and a “starry sky” pattern of hyperfluorescence on fluorescein angiography (FA) and considered an early or benign form of AMD.^7,11,17^ However, recent studies have demonstrated that cuticular drusen are a distinct phenotype of intermediate AMD^21^ and associated with development of geographic atrophy (GA),^2,16,22^ macular neovascularization (MNV),^16,23^ and acquired vitelliform lesions (AVL).^24,25^ Although one study reported that eyes with cuticular drusen did not demonstrate significant reductions in retinal sensitivity compared with eyes without cuticular drusen,^19^ a 5-year study revealed cumulative estimated risks of 28.4% for GA and 8.7% for MNV.^16^ For comparison, the Beaver Dam Eye Study found that eyes with reticular pseudodrusen had a cumulative incidence of GA of 21–36% and MNV of 13–20%, whereas eyes with soft drusen demonstrated cumulative incidences of GA of 9–11% and MNV of 8–10%.^26^ These findings underscore that cuticular drusen may carry a meaningful risk for progression to late stage AMD.

The phenotype, structural characteristics, and progression patterns of cuticular drusen have been characterized using standard clinical ophthalmic imaging, including optical coherence tomography (OCT),^7,8,10,15,22,27^ fundus autofluorescence (FAF),^7,15,28^ and fluorescein angiography (FA).^11^ However, these modalities lack the spatial resolution required to directly resolve cellular-level alterations in photoreceptors and the RPE, limiting insight into the earliest microstructural consequences of cuticular drusen. Adaptive optics (AO) has enabled retinal imaging at cellular resolution, providing a powerful tool for investigating the microstructural consequences of this distinct drusen subtype on photoreceptors and their supporting RPE.^29^ Informed by histological correlations, AO ophthalmoscopy has elucidated the phenotypic features and life cycle of other drusen types, including soft drusen^30,31^ and subretinal drusenoid deposits (SDDs; also referred to as reticular pseudodrusen).^32–36^ However, comparable studies of cuticular drusen are lacking, and their impacts on adjacent photoreceptors and RPE remain poorly understood.

Recently, cuticular drusen have been classified into distinct subtypes based on their morphology and topographic distribution using spectral domain-OCT and FAF.^15,16^ These classifications provide a foundation for applying high-resolution AO imaging to study cuticular drusen and their impact on photoreceptors and RPE. The purpose of this study was to address the knowledge gaps by providing a detailed examination of the chorioretinal structure surrounding cuticular drusen through cross-sectional analysis and longitudinal follow-up, using multimodal imaging, including adaptive optics scanning laser ophthalmoscopy (AOSLO).^32^ This study offers new insights into the structure, evolution, and photoreceptor consequences of cuticular drusen, with important implications for understanding the pathophysiology of AMD.

## Methods

The study followed the tenets of the Declaration of Helsinki, complied with the Health Insurance Portability and Accountability Act of 1996, and was approved by the Institutional Review Boards at the University of Alabama at Birmingham and the University of California – Los Angeles. Written informed consent was obtained from all participants.

This was a retrospective study using a subgroup of study patients recruited for adaptive optics imaging of AMD.^33,37^ The procedures for subject enrollment, image acquisition, and image processing have been described previously.^33,36–38^ For completeness, we provide a summary of the study methodology here.

### Subjects

Study participants were recruited from clinical research registries and retina clinics at university-affiliated ophthalmology centers. Inclusion criteria were subjects diagnosed with AMD with Snellen visual acuity of 20/200 or better. Exclusion criteria included diabetes, glaucoma, history of retinal vascular occlusions and hereditary retinal dystrophy. Refractive errors determined from the medical record were within ±6 D spherical and ±3 D cylinder for all participants.

AMD severity was further assessed by a masked, experienced grader using the Age-Related Eye Disease Study 2 (AREDS2) severity scale for AMD based on 30° color fundus photographs.^39^ Only patients with early to intermediate AMD (AREDS2 grade 2 to 8) and with predominantly cuticular drusen were included in this analysis.

### Multimodal imaging

CFP was obtained using a flash fundus camera (FF450 Plus, Carl Zeiss Meditec, Dublin, CA). Near-infrared reflectance (NIR, λ = 830 nm), FAF (excitation, 488 nm; emission > 600 nm), and OCT (λ = 870 nm; scan depth = 1.9 mm; axial resolution = 3.5 μm per pixel in tissue; lateral resolution = 14 μm per pixel in tissue) were acquired with a confocal SLO and spectral domain OCT (Spectralis HRA+OCT2; Heidelberg Engineering, Carlsbad, CA). Multimodal images were registered using custom and commercial software (Photoshop, Adobe Systems Inc., Mountain View, CA).

### AOSLO imaging

High resolution retinal images were acquired using a research AOSLO system developed in our laboratory.^40,41^ This instrument employed a low coherent superluminescent diode (HP 840, Superlum Ltd, Moscow, Russia. λ = 840 nm, Δλ = 50 nm) and acquired images with a field of view of 1.2° × 1.2° at a frame rate of 15 Hz. The subject’s pupil was dilated with 1.0% tropicamide and 2.5% phenylephrine hydrochloride before imaging. Photoreceptor images were recorded continuously over an area of approximately 15° × 15° to 20° × 20° in the macula. AOSLO images were subsequently processed to correct for the nonlinearity induced by the resonant scanning, registered to enhance signal to noise ratio, and montaged to generate a wide-field high-resolution retinal image.

CFP, NIR and FAF images were magnified to match the AOSLO scale and then aligned with the AOSLO montage using custom software.

### Cuticular drusen identification and phenotyping

On CFP, cuticular drusen appeared as numerous small, sharply demarcated round, yellow deposits. On FAF, they manifested as densely packed hypoautofluorescent focal dots with a hyperautofluorescent rim. Oncorresponding OCT B-scans, these lesions were identified as well-demarcated elevations of the RPE-BrM complex with a characteristic sawtooth configuration accompanied by distinctive posterior hypertransmission stripes extending into the choroid.^11,15–17^ Eyes demonstrating at least 50 bilaterally distributed lesions with these multimodal imaging features were considered having cuticular drusen.^7,42^

OCT served as the primary modality for lesion identification. Lesions were classified as cuticular drusen only if they met cuticular morphology criteria on OCT B-scans and demonstrated characteristic posterior hypertransmission into the choroid. En face imaging, including en face OCT, was used to confirm lesion spatial distribution, assess areal coverage, and estimate lesion number. An eye was considered as predominantly cuticular drusen when ≥80% of all drusen within the central 6-mm retina (macula) met these criteria and accounted for the largest cumulative drusen area, with no dominant soft drusen or subretinal drusenoid deposits present.

The identified cuticular drusen were classified into three subtypes based on OCT cross-sectional structure.^15^ Type 1 lesions exhibited shallow elevation of the RPE basal lamina without clear discernible internal content. Type 2 exhibited triangular elevations of the RPE resulting in a characteristic sawtooth appearance with hyporeflective internal contents. Type 3 exhibited broad, mound shaped elevations of the RPE band with hyporeflective internal contents.

### Image analysis

Using the registered CFP, NIR, and FAF images, cuticular drusen appearing on the clinical standard imaging were localized on AOSLO images, which were further examined and confirmed by inspection of corresponding OCT B-scans. The impact of cuticular drusen on the overing retina was evaluated by high resolution AOSLO. ImageJ (Version 1.54i 03 March 2024, https://imagej.nih.gov/ij/) was used to calculate the quasi-circular areas of cuticular drusen evident in the AOSLO images, following previously reported methods for assessing lesion-affected retinal regions.^34,36^ Specifically, the equivalent diameter *d_i_* (µm) was calculated as 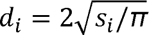, where *s_i_* is the retinal area affected by an individual lesion, and π is the constant ratio of a circle’s circumference to its diameter. To assist manual delineation, the border of the hyporeflective region associated with each lesion was identified and enhanced using a standardized 2-step processing procedure (Photoshop, Adobe Systems Inc., Mountain View, CA). The image was first filtered (Filter > Blur > Gaussian Blur. Radius = 3 pixels), then sharpened (Filter > Sharpen > Smart Sharpen. Amount = 180%, Radius = 64 pixels).^34,36^

## Results

Nineteen eyes (OD, 11; OS, 8) from 12 patients were studied (**Table 1**). Subject age was 70.3 ± 5.8 years. The cohort included 5 males and 7 females. AMD disease severity, based on the AREDS2 grading step, ranged from step 2 to step 7. Visual acuity was 0.15 ± 0.17 in the right eye (OD) and 0.04 ± 0.16 logMAR in the left eye (OS).

**Table 1.** Study Cohort Characteristics.

| Characteristic | Value |
| --- | --- |
| Number of patients | 12 |
| Sex (male/female) | 5/7 |
| Age, years (mean ± SD) | $70.3 \pm 5.8$ |
| Age range, years | 62 - 78 |
| AREDS2 grade | 2 - 7 |
| Visual acuity (logMAR) |  |
| OD | $0.15 \pm 0.17$ |
| OS | $0.04 \pm 0.16$ |
| Total eyes analyzed | 19 |

Of the 19 eyes examined, 17 demonstrated cuticular drusen only, 1 eye showed a few coexisting soft, confluent drusen in the macula, and 1 eye was accompanied by acquired vitelliform lesion. Six eyes of 3 subjects were longitudinally followed for durations of 20, 44, and 47 months, respectively.

A total of 3,177 cuticular drusen were analyzed across 19 study eyes. Of these lesions, 2,570 (80.9%) cuticular drusen were classified by OCT as Type 1, 550 (17.3%) Type 2, and 57 (1.8%) Type 3. The mean diameters differed significantly among types, measuring 52.62 ± 9.38 µm for Type 1, 71.98 ± 12.39 µm for Type 2, and 124.72 ± 20.94 µm for Type 3 (**Table 2**).

**Table 2.** Lesion characteristics.

| Type | Number | Size [Mean ± SD (Min - Max) (μm)] |
| --- | --- | --- |
| Type 1 | 2570 (80.9%) | 52.62 ± 9.38 (27.07 – 86.72) |
| Type 2 | 550 (17.3%) | 71.88 ± 12.39 (44.87 – 115.84) |
| Type 3 | 57 (1.8%) | 124.72 ± 20.94 (80.90 – 161.33) |

High-resolution AOSLO combined with multimodal imaging revealed phenotype-specific disruption of photoreceptor and RPE optical properties associated with cuticular drusen. As a reference for the normal retinal appearance across imaging modalities, **Figure 1** presents multimodal images of a healthy retina. CFP shows a normal fundus without drusen or pigmentary abnormalities, FAF demonstrates natural variation in fundus autofluorescence, and spectral-domain OCT displays continuous, well-aligned outer retinal hyperreflective bands, including the external limiting membrane (ELM), ellipsoid zone (EZ), interdigitation zone (IZ), and the RPE–Bruch’s membrane complex. AOSLO renders a clearly resolved photoreceptor mosaic (magnified boxes), with variability in individual photoreceptor brightness consistent with known dynamic reflectance properties of cones and rods.^43,44^

**Figure 1.**
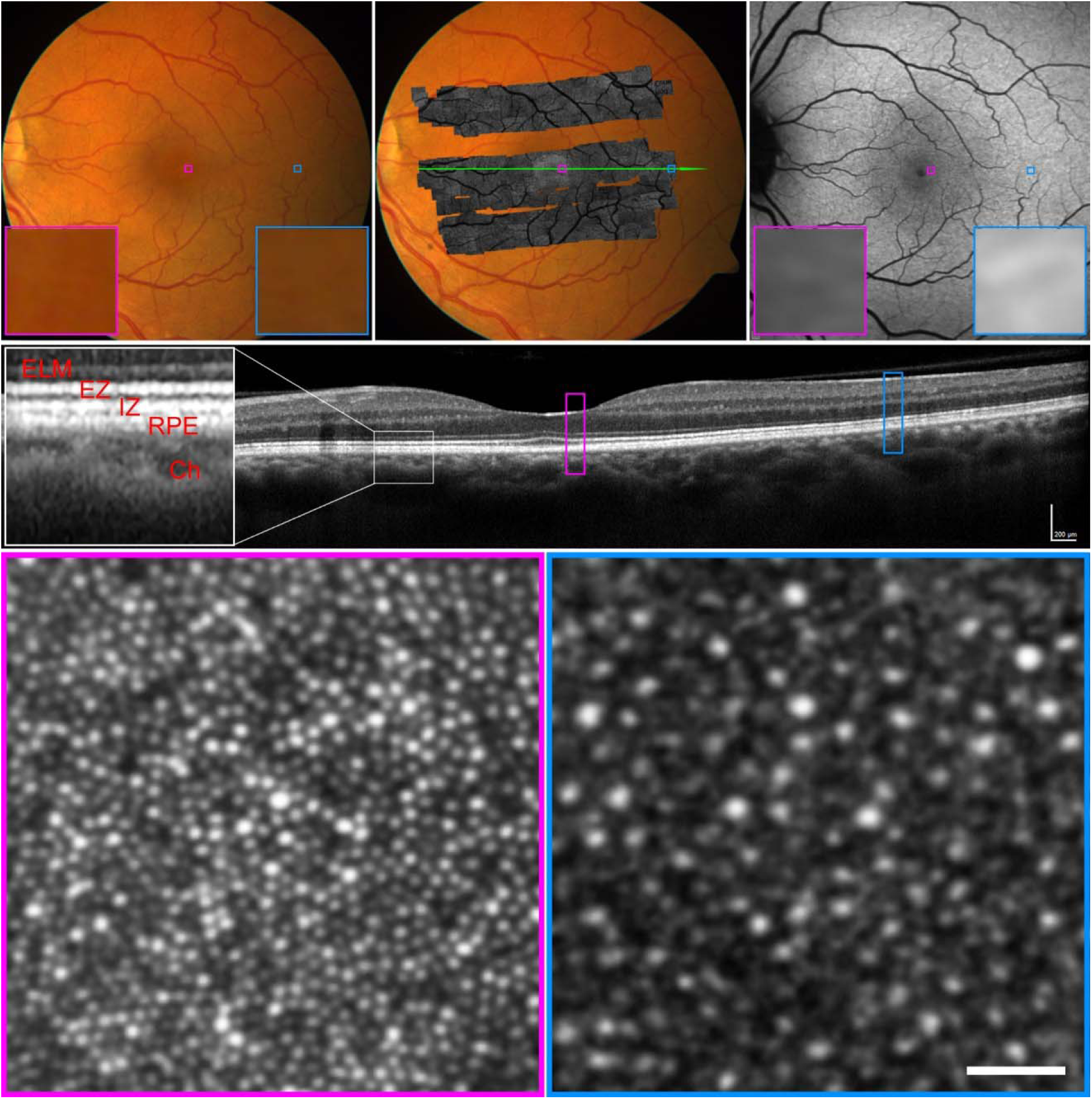
Multimodal imaging of a healthy retina. Top left: Color fundus photography (CFP) of 30° field. Top middle: Adaptive optics scanning laser ophthalmoscopy (AOSLO) montage overlaid on the CFP. Top right: Fundus autofluorescence (FAF) of 30° field. Middle row: Optical coherence tomography (OCT) B-scan taken along the green line in the top middle panel, showing uniformly reflective outer retinal bands, including the external limiting membrane (ELM), the ellipsoidal zone (EZ), the interdigitation zone (IZ), and the retinal pigment epithelium (RPE)-Bruch complex (magnified white box). Bottom: AOSLO images corresponding to the magenta and blue boxed regions in the OCT B-scan. Bottom left: Photoreceptor mosaic at 0.7° eccentricity, where bright punctate dots represent cones. Bottom right: Photoreceptor mosaic at 9.3° eccentricity showing cones (larger bright dots) and rods (smaller dim dots). Reflectivity varies among individual photoreceptors.^43^ Images were acquired from the right eye of subject ACADIA-019 (Age-Related Eye Disease Study grade 1). Scale bars: 100 _μ_m (OCT), 25 _μ_m (AOSLO).

AOSLO imaging of Type 1 cuticular drusen (**Figure 2**) revealed localized perturbation of photoreceptor reflectivity. Most cones remained visible and retained reflectance over lesions but situated on a markedly hyporeflective background (**Figure 2** bottom panel, color arrowheads) relative to surrounding regions. With Type 2 cuticular drusen (**Figure 3**), AOSLO showed more pronounced cone photoreceptor disruption than in Type 1 lesions. Cones overlying these lesions exhibited reduced reflectivity, and in some regions individual photoreceptors lost reflectivity entirely, resulting in larger hyporeflective patches. AOSLO imaging of Type 3 cuticular drusen (**Figure 4**) illustrated substantial photoreceptor disruption over these lesions. The cone mosaic disappeared and retinal areas over lesions exhibited pronounced hyporeflectivity that exceeded those observed in Types 1 and 2 lesions. The effects of cuticular drusen on photoreceptor structure and retinal reflectivity, as described above, are collectively illustrated in **Figure 5**. AOSLO provided microscopic en face visualization of cuticular drusen morphology, revealing circular, oval, lobular, and multilobulated patterns.

**Figure 2.**
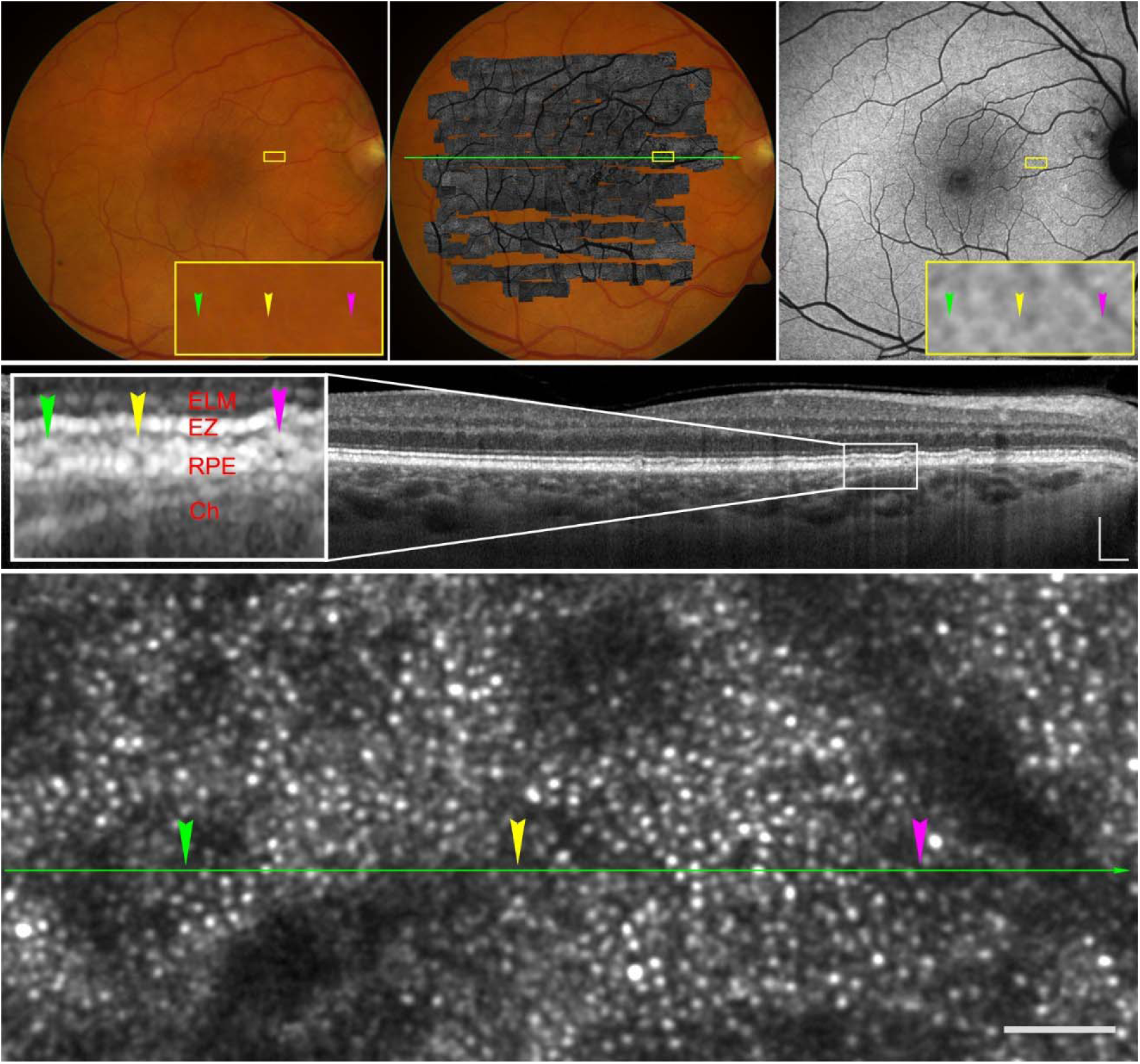
Multimodal imaging of Type 1 cuticular drusen. Top left: Color fundus photography (CFP) of a 30° field. The magnified yellow box shows an area containing three Type 1 cuticular drusen, indicated by color arrowheads that correspond across all imaging modalities. Top middle: Montaged adaptive optics scanning laser ophthalmoscopy (AOSLO) image overlaid on the CFP. Top right: Fundus autofluorescence (FAF) image. Type 1 lesions appear as hypoautofluorescent foci with a hyperautofluorescent rim. Middle: Optical coherence tomography (OCT) B-scan along the green line in the top middle panel, showing shallow RPE elevations with internal hyporeflectivity and associated choroidal hypertransmission. The enlarged white box corresponds to the yellow-boxed region above and highlights the Type 1 lesions. Bottom: AOSLO of the yellow-boxed region, with color arrowheads indicating the same three lesion sites. AOSLO revealed localized perturbation of photoreceptor reflectivity. Most cones remained visible and retained reflectance over lesions but situated on a markedly hyporeflective background (color arrowheads) relative to surrounding regions. Images were acquired from the right eye of subject ACADIA-036 (Age-Related Eye Disease Study 2 grade 4). Scale bars: 200 μm (OCT), 50 μm (AOSLO).

**Figure 3.**
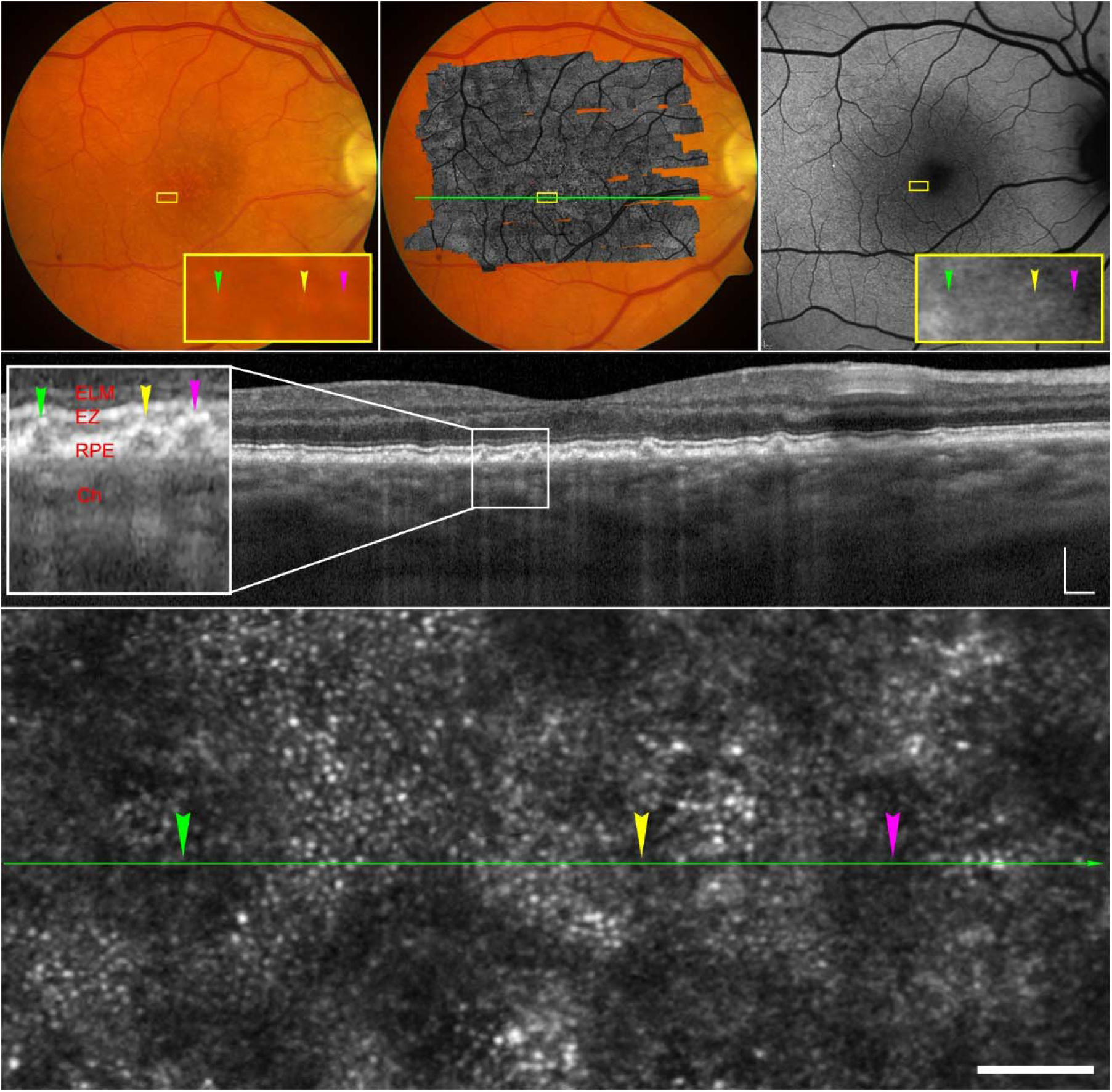
Multimodal imaging of Type 2 cuticular drusen. Top left: Color fundus photography (CFP) of a 30° field. Color arrowheads denote three Type 2 cuticular drusen across all modalities. Top middle: Adaptive optics scanning laser ophthalmoscopy (AOSLO) overlaid on the CFP. Top right: Fundus autofluorescence (FAF) of the same region. Middle: Optical coherence tomography (OCT) B-scan obtained along the green line in the top middle panel. The enlarged white box corresponds to the yellow-boxed region above and highlights Type 2 lesions. Bottom: AOSLO of the yellow-boxed region, showing more pronounced cone photoreceptor disruption than in Type 1 lesions (Figure 1). Cones overlying these lesions exhibited reduced reflectivity, and in some regions individual photoreceptors lost reflectivity entirely, resulting in larger hyporeflective patches (yellow and magenta arrowheads). Images were acquired from the right eye of subject ACADIA-029 (Age-Related Eye Disease Study 2 grade 6). Scale bars: 200 μm (OCT), 50 μm (AOSLO).

**Figure 4.**
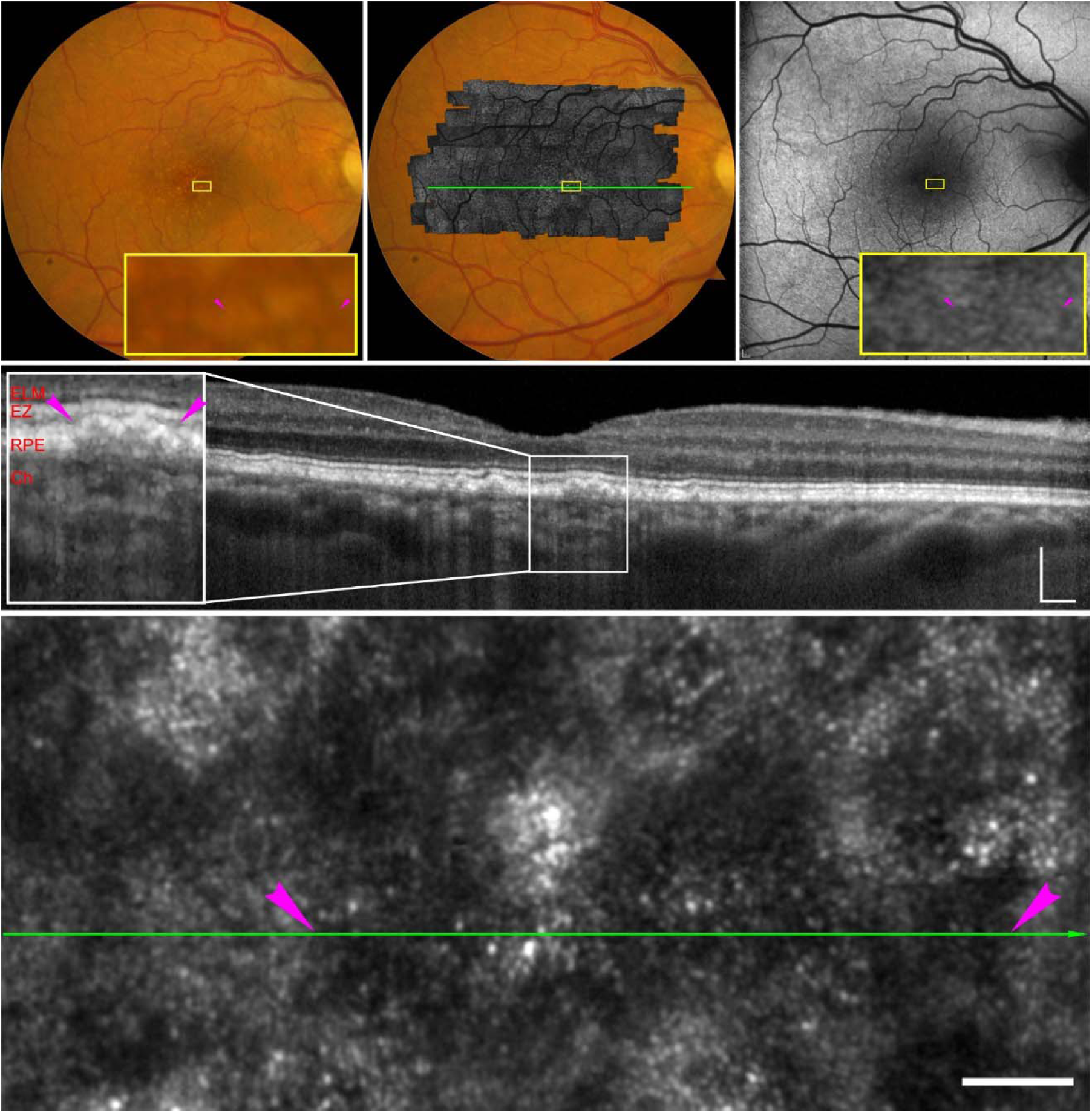
Multimodal imaging of Type 3 cuticular drusen. Top left: Color fundus photography (CFP) of a 30° field. Color arrowheads denote three Type-3 cuticular drusen across all modalities. Top middle: Montaged adaptive optics scanning laser ophthalmoscopy (AOSLO) image overlaid on the CFP. Top right: Fundus autofluorescence (FAF) of the same region. Middle: Optical coherence tomography (OCT) B-scan obtained along the green line in the top middle panel. The enlarged white box corresponds to the yellow-boxed region above and highlights Type 3 lesions. Bottom: AOSLO of the yellow-boxed region, with color-coded arrowheads marking the positions of the Type 3 drusen across all modalities. Substantial photoreceptor disruption occurred over these lesions. Cone mosaic disappeared and retinal areas over lesions exhibited pronounced hyporeflectivity that exceeded those observed in Types 1 and 2 lesions. Images were acquired from the right eye of subject AMD-001 (Age-Related Eye Disease Study 2 grade 6). Scale bars: 200 μm (OCT), 50 μm (AOSLO).

**Figure 5.**
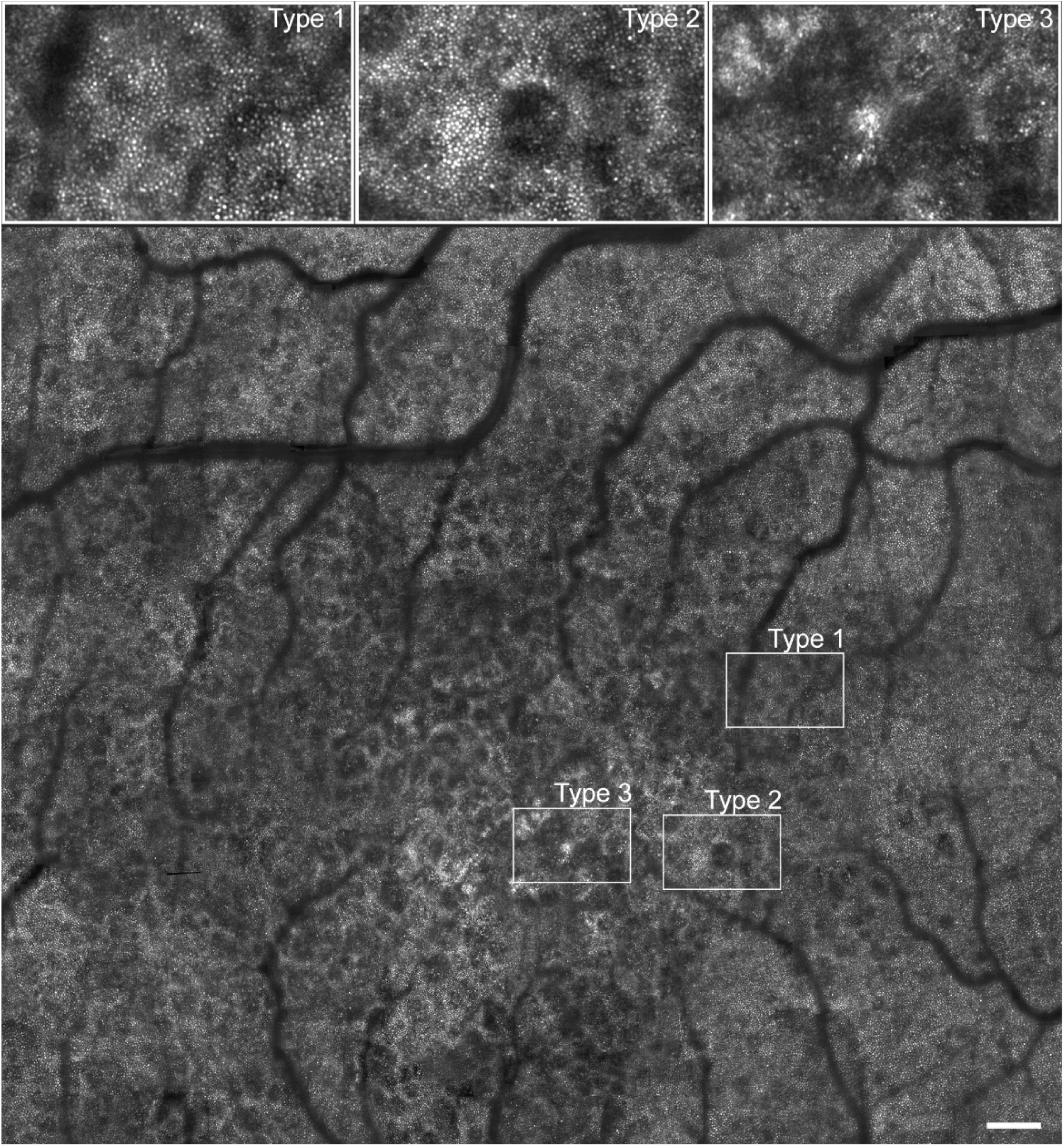
Adaptive optics scanning laser ophthalmoscopy (AOSLO) of cuticular drusen. A large AOSLO image (13.5° × 11.7° macular field) demonstrates multiple cuticular drusen with varying morphologies. Over Type 1 cuticular drusen, most cones remained visible and retained reflectance but situated on a hyporeflective background relative to surrounding regions. With Type 2 lesion, Cones exhibited pronounced reduction in reflectivity, and in some regions individual photoreceptors lost reflectivity entirely, resulting in larger hyporeflective patches. In Type 3 cuticular drusen, the cone mosaic disappeared and retinal areas over the lesions exhibited pronounced hyporeflectivity that exceeded those observed in Types 1 and 2 lesions. Images were acquired from the right eye of subject AMD-001. Scale bar: 200 μm.

Clinical standard imaging demonstrated typical appearances of cuticular drusen across phenotypes on CFP, FAF, and OCT, consistent with previous reports. Notably, OCT consistently revealed prominent choroidal hypertransmission beneath all cuticular drusen in this cohort (**Figures 2-4**).

Longitudinal follow-up demonstrated reduced retinal reflectivity on AOSLO preceding OCT detectable Type 1 cuticular drusen. On OCT, newly developed Type 1 cuticular drusen were identified at the 37-month follow-up, enabling retrospective assessment of the corresponding retinal locations on earlier imaging (**Figure 6**). Follow-up OCT disclosed multiple Type 1 cuticular drusen, characterized by shallow RPE elevations with underlying choroidal hypertransmission, while AOSLO at these locations demonstrated reduced photoreceptor and background reflectivity. Retrospective review of the earlier image at the same retinal sites revealed localized reductions in AOSLO reflectivity (**Figure 6**, color arrowheads), with photoreceptors remaining discernible on a darkened background. At these locations, baseline OCT B-scans (outlined by colored boxes) showed intact outer retinal bands, including the ELM, EZ, IZ, and RPE, without discernable RPE elevation, choroidal hypertransmission, or cuticular drusen.

**Figure 6.**
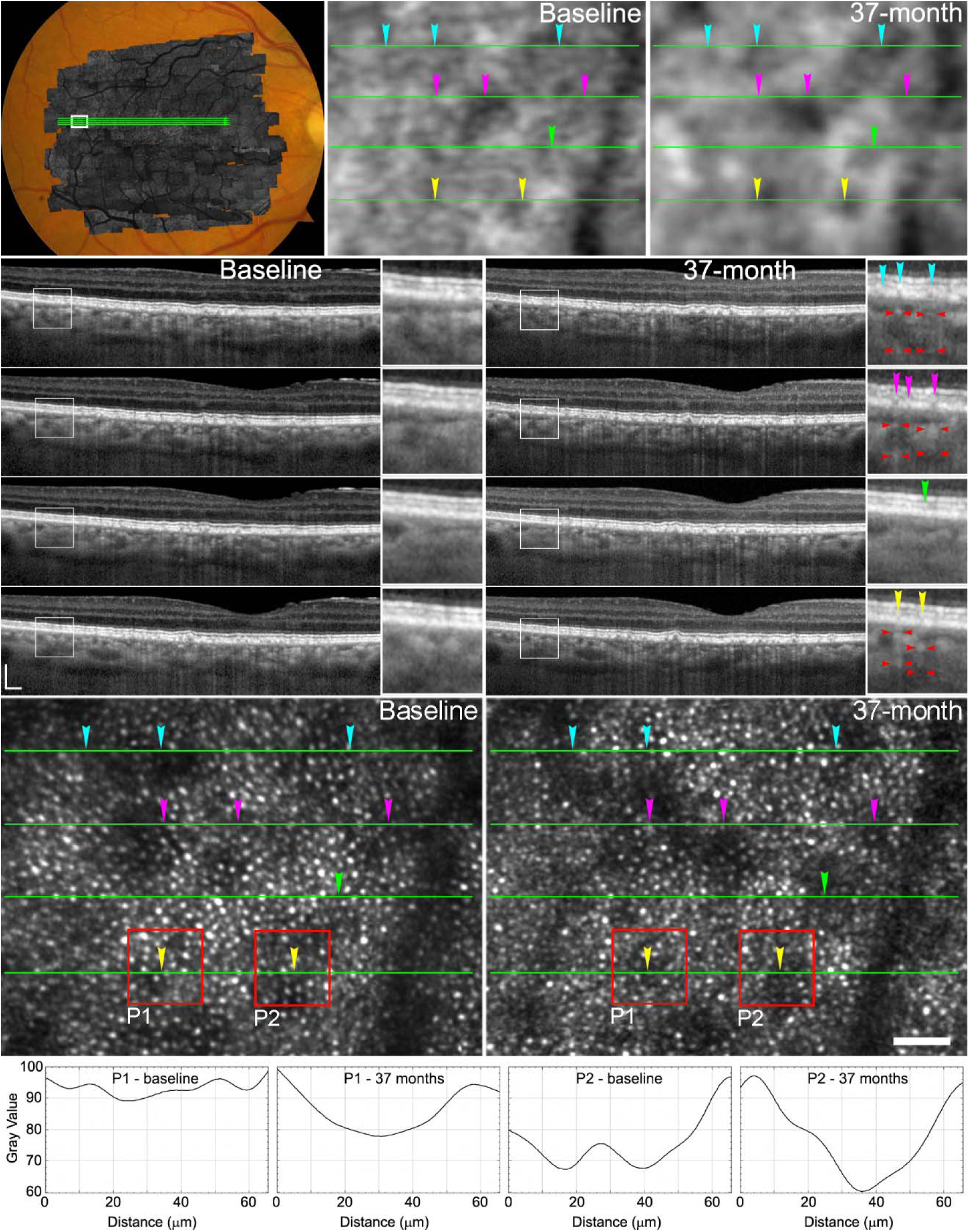
Adaptive optics scanning laser ophthalmoscopy (AOSLO) and optical coherence tomography (OCT) of new cuticular drusen development. Top row: AOSLO montage overlaid on color fundus photography (CFP). The white box delineates a region in which new cuticular drusen developed during follow-up; green lines indicate the locations of four consecutive OCT B-scans. Fundus autofluorescence (FAF) images of the boxed region in the CFP at baseline and 37-month follow-up; arrowheads mark lesion sites. Second to fifth rows: Consecutive OCT B-scans at baseline (left) and follow-up (right). At baseline, all marked regions show intact outer retinal bands (ELM, EZ, IZ, and RPE) without choroidal hypertransmission. At follow-up, most regions demonstrate newly formed Type 1 cuticular drusen, characterized by shallow RPE elevation and associated choroidal hypertransmission (red arrowheads). Sixth row: Corresponding AOSLO images. At baseline, photoreceptors exhibit preserved reflectivity on a hyporeflective background. At follow-up, photoreceptor reflectivity is reduced on a more pronounced hyporeflective background. Red boxes highlight two newly developed lesions confirmed on OCT. Bottom row: Reflectivity profiles from the red boxed regions show reduced trough-to-edge reflectivity ratios (P1: 0.91 to 0.80; P2: 0.77 to 0.63). Images were acquired from the right eye of subject AMD-001. ELM: external limiting membrane; EZ: ellipsoid zone; IZ: interdigitation zone; RPE: retinal pigment epithelium. Scale bars: 200 μm (OCT), 50 μm (AOSLO).

Focal retinal reflectivity reductions detected by AOSLO at sites of subsequent cuticular drusen formation were also confirmed on FAF (**Figure 6**, top row). Careful inspection of baseline FAF images at locations corresponding to AOSLO abnormalities revealed subtle hypoautofluorescence in regions that later developed cuticular drusen, which became more pronounced on follow-up imaging. These findings corroborate that AOSLO-detected retinal reflectivity abnormalities precede OCT-visible structural changes. By retrospectively tracing newly formed lesions to their baseline locations, hyporeflective regions as small as 15 µm were identified before lesions became clinically discernible.

The continuum of morphologic progression of cuticular drusen was also demonstrated by longitudinal OCT (**Supplementary Figure 1**), showing that Type 3 lesions evolved from the coalescence and enlargement of Type 2 cuticular drusen over time. These results corroborate findings from previous studies.^15,20^

## Discussion

Using multimodal imaging with cellular-resolution AOSLO, we identified phenotype-dependent alterations in photoreceptor and RPE optical properties associated with cuticular drusen in eyes with AMD. Cone reflectivity changes varied systematically across OCT-defined lesion types and were most pronounced in more advanced phenotypes. Whereas Type 1 cuticular drusen were associated with a detectable, albeit slightly disrupted cone mosaic. The cone reflectivity was attenuated with Type 2 cuticular drusen and altogether absent in association with Type 3 lesion.

Notably, focal reductions in outer retinal reflectivity on AOSLO preceded OCT-detectable lesion formation, whereas choroidal hypertransmission was consistently observed across lesion types. These findings collectively underscore the complementary roles of AOSLO and OCT in delineating the microstructural continuum of cuticular drusen and provide insight into the earliest detectable events in lesion formation. In contrast to prior work largely focused on late-stage AMD,^14,16^ our study emphasizes early-stage cuticular drusen, offering an opportunity to identify sensitive structural biomarkers that may inform risk stratification and earlier intervention strategies.

Foundational mechanistic work by Spaide and Curcio modeled perturbations of chorioretinal optical properties associated with cuticular drusen and explained their appearance on multimodal imaging.^9^ Balaratnasingam and colleagues subsequently refined the clinical and histologic characterization of cuticular drusen and delineated their subtypes and life cycle using spectral-domain OCT.^15^ Building on these prior frameworks, our study extends these observations by providing cellular-level in vivo assessment of photoreceptor alterations, revealing a gradation of cone reflectivity changes across cuticular drusen phenotypes or stages, with relative preservation over Type 1 lesions and progressively reduced or absent reflectivity over Type 2 and Type 3 lesions (**Figures 2–6**).

These outer retinal reflectivity changes paralleled OCT-defined structural alterations of the RPE, supporting a close relationship between lesion maturation and photoreceptor optical disruption. Because cone reflectivity in confocal AOSLO depends on outer segment integrity and directional waveguiding,^45^ these changes reflect disrupted or impaired waveguiding and early functional compromise of the photoreceptors, which may contribute to localized reductions in visual sensitivity.^25,46,47^

Precise lesion size measurement is important for assessing cuticular drusen progression and their impact on chorioretinal structure and function. High-resolution AOSLO enables microscopic visualization of cuticular drusen and their effects on overlying photoreceptors and the RPE, complementing OCT, which may be limited by lateral resolution and en face scan spacing when evaluating small lesions.^11,21^ AOSLO-based lesion size measurements, derived from the en face area of affected retina serving as a surrogate for lesion dimensions, were consistent with prior reports.^11,20,21,27,48^ Using AOSLO, we identified nascent cuticular drusen as small as 15 µm (**Figure 5**), approaching histologic measurements (11.87 µm),^20^ highlighting the sensitivity of this modality for early lesion detection.

A key longitudinal finding was that AOSLO revealed focal outer retinal hyporeflectivity at locations that later developed overt cuticular drusen on OCT (**Figure 6**). These hyporeflective regions preceded OCT-detectable lesions, with cones remaining visible against a hyporeflective background and choroidal hypertransmission emerging only after lesion appearance. This temporal sequence suggests early alterations in the optical properties of the photoreceptor–RPE complex before overt structural lesion formation. The observed reflectivity reductions may reflect increased light transmission to the choroid due to changes in the photoreceptor–RPE–choroid complex or RPE-intrinsic alterations, such as altered melanin distribution or melanosome organization,^9^ while photoreceptor reflectivity itself remains preserved due to minimal architectural impact. Although the underlying cellular mechanisms remain uncertain, the observations are consistent with impaired RPE light attenuation^9^ occurring early in the cuticular drusen life cycle and highlight important targets for future investigation.

Choroidal hypertransmission was consistently observed beneath newly developed lesions, reflecting increased light transmission through the photoreceptor-RPE complex into the choroid. This finding is consistent with the Spaide–Curcio model,^9^ and characteristic donut-pattern reversal recently described.^21^

Choroidal hypertransmission has been a recognized consequence of RPE atrophy.^28,35^ However, the temporal pattern of hypertransmission associated with cuticular drusen differs markedly from that seen with subretinal drusenoid deposits (SDD).^35^ SDD associated choroidal hypertransmission appeared only in advanced or regressing stages of the lesions. The much earlier onset of hypertransmission associated with cuticular drusen underscores fundamental differences in the pathophysiology of these lesions at the photoreceptor–RPE–Bruch’s membrane complex. FAF findings further support early RPE involvement, demonstrating darkened retinal areas with associated choroidal hypertransmission prior to overt lesion appearance on OCT (**Figure 6**). These observations are in line with findings from prior imaging and histologic studies that demonstrated incomplete RPE coverage over nodular and cuticular drusen.,^15,20,49,50^ supporting the concept that early pathologic events originate within the RPE.^25^

The strengths of this study include the use of registered multimodal imaging, enhanced by high resolution AOSLO in a well-defined cohort of subjects with predominantly cuticular drusen. Cross-sectional and longitudinal follow-up assessment in a large sample of individual lesions enabled detailed analysis of lesion-specific changes. The high lateral resolution of AOSLO (2.8 μm, ∼1/10th the diameter of smallest clinically measurable cuticular drusen^11^), and the use of the photoreceptor mosaic as an en face reference allowed precise localization of lesion effects. In addition, accurate alignment of OCT with AOSLO provided stage-specific characterization of lesion structure and associated alterations in overlying photoreceptors and RPE with greater accuracy than either modality alone. Together, these features enabled a unique and detailed evaluation of the cuticular drusen phenotype in early and intermediate AMD.

This study has several limitations. The sample size was small, and functional correlative data was not available, limiting our ability to relate structural alterations to visual performance. Thus, the findings pertain only to local photoreceptor perturbation and should not be interpreted as having direct prognostic significance. Our AOSLO imaging was confined to the macular region and did not evaluate the more peripherally distributed cuticular drusen described in larger cohort studies, such as those by Balaratnasingam and co-workers.^15^ Moreover, the limited number of subjects and relatively short follow-up period were insufficient to document the full evolutionary spectrum of cuticular drusen, including their enlargement, coalescence into soft drusen, or progression to atrophy or neovascularization. Longer-term imaging will be essential to establish the complete natural history of this phenotype. Another limitation of this study relates to the definitive differentiation of cuticular drusen from soft drusen. FA, and more recently en face OCT,^21^ can improve the reliability of differentiation but these modalities were not available in this study. FA was not justified in this intermediate AMD cohort because of its risks and the lack of clinical indication. Prior work by Høeg and colleagues demonstrated substantial agreement between color fundus photography and FA for detecting cuticular drusen,^13^ and Balaratnasingam and co-workers^15^, Leng and colleagues^8^, have provided clear multimodal imaging criteria, especially using OCT, for reliable identification. Accordingly, we used multimodal imaging to ensure confident detection of cuticular drusen in this study.

In conclusion, multimodal imaging with cellular resolution reveals phenotype-dependent alterations in photoreceptor- and RPE-associated optical properties overlying cuticular drusen in AMD. Longitudinal AOSLO imaging demonstrated that localized reductions in outer retinal reflectivity precede OCT-detectable lesion formation, indicating that microscopic alterations in outer retinal and RPE optical properties occur early in cuticular drusen life cycle. These findings highlight the complementary value of AOSLO and OCT for delineating the structural continuum of cuticular drusen and support the utility of high-resolution in vivo imaging for defining disease progression and the natural history of this AMD phenotype.

## Declaration

This manuscript describes original work that has not been published or submitted elsewhere. A revised version of this work will be submitted to American Journal of Ophthalmology.

## Ethics

The study followed the tenets of the Declaration of Helsinki, complied with the Health Insurance Portability and Accountability Act of 1996, and was approved by the Institutional Review Boards at the University of California at Los Angeles. Written informed consent was obtained from all participants.

## Data Availability

All data produced in the present work are contained in the manuscript.

## Acknowledgments

The authors thank Christine A Curcio, PhD, Cynthia Owsley, PhD, C. Douglas Witherspoon, MD, Christopher A Girkin, MD, and Mark E Clark, BS, for helping with data acquisition.

---

**Supplement Figure 1.**
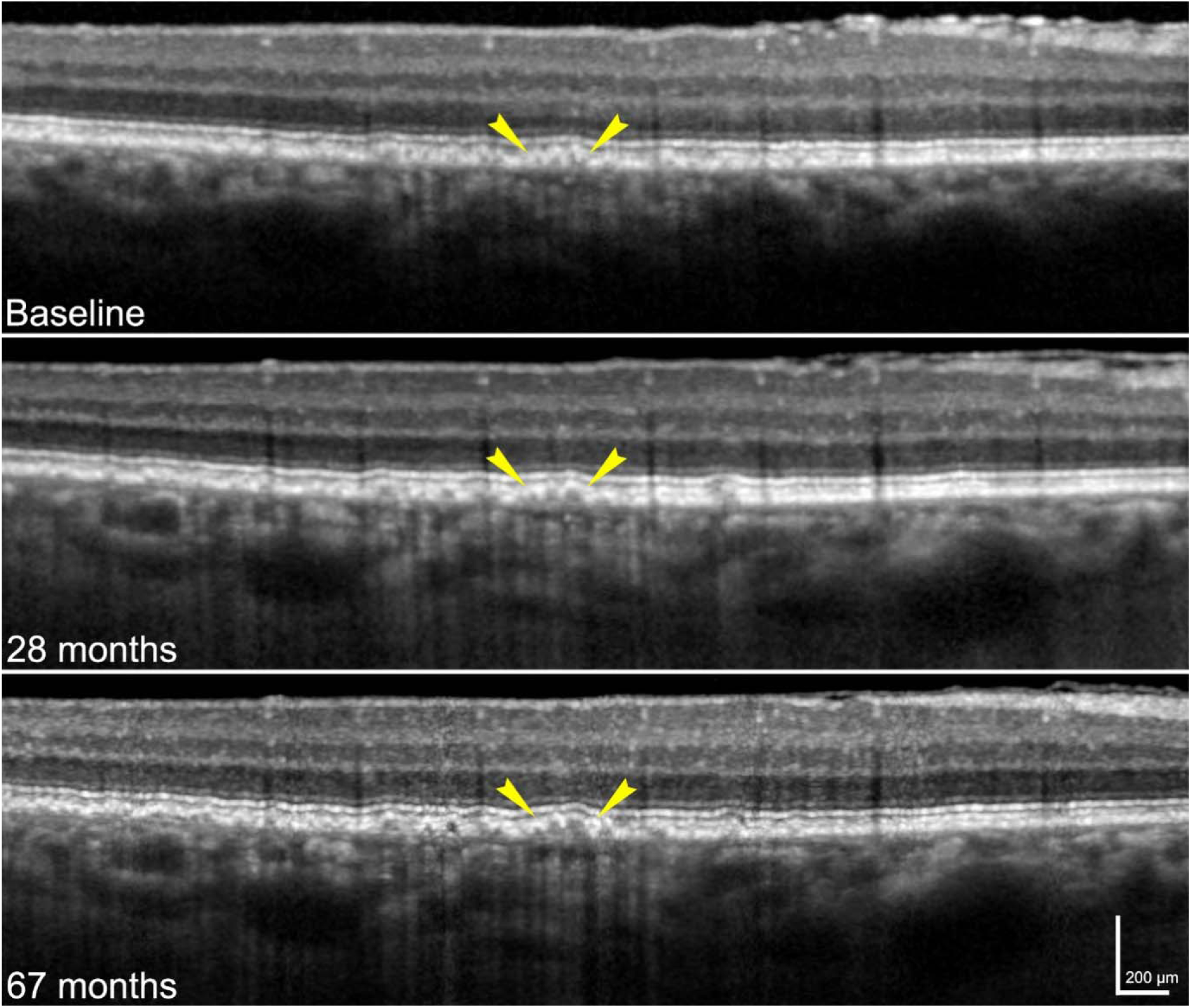
Optical coherence tomography of cuticular drusen progression. Regions marked by yellow arrowheads show Type 1 cuticular drusen at baseline that progressed to Type 2 by 28-month and then coalesce into Type 3 by 67-month. Images were acquired from the eye of subject AMD-001.

